# COVID-19 in 823 Transplant patients: A Systematic Scoping Review

**DOI:** 10.1101/2021.01.18.21250025

**Authors:** Moataz Maher Emara, Mahmoud Elsedeiq, Mohamed Elmorshedi, Hamed Neamatallah, Mostafa Abdelkhalek, Amr Yassen, Ashraf Nabhan

**Affiliations:** Department of Anesthesiology and Intensive Care and Pain Medicine, Mansoura University, Egypt; Liver Transplantation Program, Gastrointestinal Surgical Center, Mansoura University, Egypt; Department of Obstetrics and Gynecology, Faculty of Medicine, Ain Shams University, Cairo, Egypt; Egyptian Center for Evidence Based Medicine, Cairo, Egypt

**Keywords:** Organ Transplant, Coronavirus, COVID-19, Systematic scoping review

## Abstract

**Background:** Management of COVID-19 in transplant patients is a big challenge. Data on immunosuppression management, clinical picture, and outcomes are lacking.

**Objectives:** To summarize the current literature on COVID-19 in transplant patients especially the data regarding the immunosuppression protocols, clinical presentation, and outcomes.

**Search strategy:** A systematic search of MEDLINE, EBSCO, CENTRAL, CINAHL, LitCovid, Web of Science, and Scopus electronic databases. The references of the relevant studies were also searched. The search was last updated on June 3, 2020.

**Selection Criteria:** Primary reports of solid organ transplant patients who developed COVID-19. An overlap of cases in different reports was checked.

**Data collection and analysis:** A descriptive summary of immunosuppression therapy (before and after COVID-19), clinical presentation (symptoms, imaging, laboratory, and disease severity), management (oxygen therapy, antiviral, and antibacterial), major outcomes (Intensive care admission, invasive mechanical ventilation, acute kidney injury), and mortality.

**Main results:** We identified 74 studies reporting 823 cases of solid organ transplantation with COVID-19. Among 372 patients, 114 (30.6%) were mild COVID-19, 101 (27.2%) moderate, and 157 (42.2%) severe or critical.

Major outcomes included intensive care unit admission, invasive ventilation, and acute kidney injury, which occurred in 121 (14.7%), 97 (11.8%), and 63 (7.7%) of patients, respectively. Mortality was reported in 160 (19.4%) patients. Missing individual data hindered making clinical correlations.

**Conclusion:** COVID-19 in solid organ transplant patients probably has a more disease severity, worse major outcomes (Intensive care admission, invasive ventilation, acute kidney injury), and higher mortality than in non-transplant patients.

## Introduction

Severe acute respiratory syndrome coronavirus-2 (SARS-CoV-2) causes the clinical syndrome called COVID-19. Infections in organ transplant patients are of special concern due to lifelong immunosuppression, common comorbidities, and the effects of some immune suppressants (diabetogenesis, neutrocytopenia, or lymphopenia).^1^ Reports are controversial regarding the clinical presentation and outcomes in those patients in comparison to the general population.

The ideal management of immunosuppressive therapy during COVID-19 in transplant patients is unclear. The balance between the increased risk of infections and graft rejection is vital. Theoretically, immunosuppression may reduce the cytokine storm syndrome - a major pathology in COVID-19 - and calcineurin inhibitors (CNI) reduce in vitro viral replication.^2^

The lack of data in the time of the COVID-19 pandemic pushed us to systemically review and summarize the available knowledge of COVID-19 in transplant patients, especially as regards the immunosuppressive management, clinical presentation, and major outcomes (admission to Intensive Care Unit (ICU), invasive mechanical ventilation (MV), acute kidney injury (AKI)), and mortality.

## Material and Methods

This review followed the Arksey and O’Malley framework for scoping reviews and the Preferred Reporting Items for Systematic Review and Meta-Analysis (PRISMA) – extension for scoping review.^3,4^

### I- Identifying research questions

Our main research questions were: (1) What is the common immunosuppressive protocol in transplant patients with COVID-19?; (2) What is the clinical presentation and disease severity in transplant patients with COVID-19?; (3) What are the major outcomes including the incidence of ICU admission, Invasive MV, AKI, and mortality?

### II- Identifying relevant studies

A systematic search in MEDLINE, EBSCO, CENTRAL, CINAHL, LitCovid, Web of Science, and Scopus electronic databases was conducted with no language restrictions. The search was last updated on June 3, 2020.

We searched the list of references for the selected study and contacted authors of published reports for additional information. The detailed search strategy is supplemented in the file (S1).

### III- Study selection

Two authors (MME and MEs) independently screened the titles and abstracts of the primary search results, then reviewed the full relevant articles. We included any article reporting original research on organ transplant patients with COVID-19.

The diagnosis of COVID-19 was considered either by clinical, radiological, or reverse-transcription polymerase chain reaction (rt-PCR). We reported case severity as reported in the primary studies.

If MME and MEs could not agree on the inclusion of any study, a third reviewer’s opinion (AY or AN) was asked.

### IV- Data charting

A data-charting electronic sheet was developed by MME and revised by all authors. Two authors (MEs and HN) independently extracted data and (MEm and MA) continuously updated the datasheet. Then, the data was collated by MME.

We extracted the following data: (1) General data (title, year of publication, authors, and country); (2) Methodological data (study design, sample, and patient characteristics - e.g. age, the transplanted organ, from living or deceased donor, duration since transplantation); (3) Immunosuppressive therapy (before and after COVID-19); (4) Clinical data (clinical presentation, imaging, laboratory investigations, and disease severity); (5) Management including (oxygen therapy, antiviral, antibacterial, mechanical ventilation); (6) Outcomes (ICU admission, Invasive MV, AKI, mortality).

This scoping review did not include a critical appraisal of the primary studies.

### V- Summarizing results

We organized our results in categories: immunosuppression, the clinical presentation including severity, and major outcomes. We presented the results as number (%) based on the available data.

## Results

We identified 74 primary studies reporting 823 organ transplant cases who developed COVID-19 after organ transplantation. (Figure 1) charts the process of studies’ inclusion.

**Figure 1.**
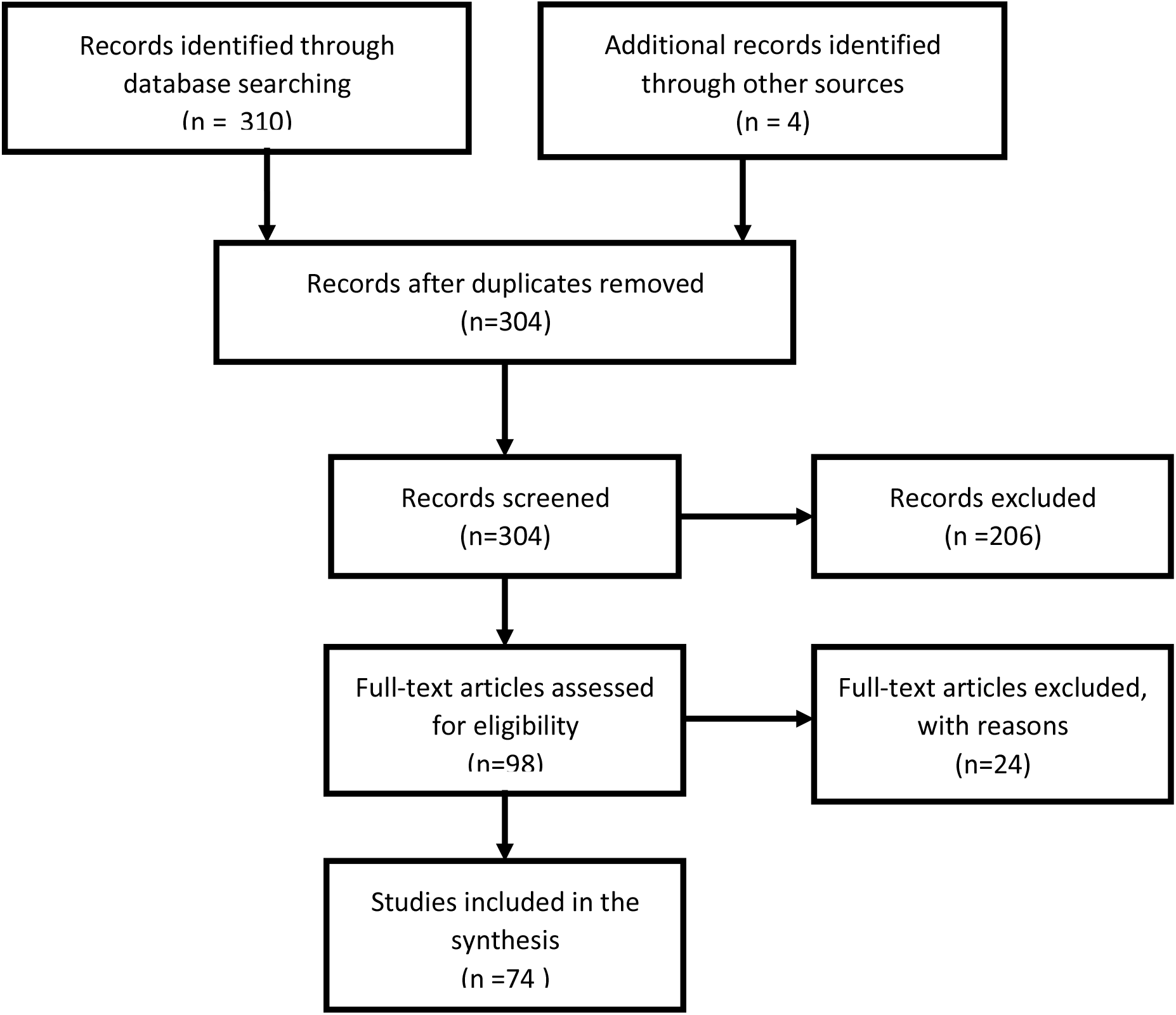
Study flowchart

These reports included 617 kidney, 98 liver, 59 heart, 31 lung, 17 combined-organ transplants, and 1 pancreas.

We tabled the studies according to the study design, country, and the number of cases (Table 1).^5-78^

**Table 1.**
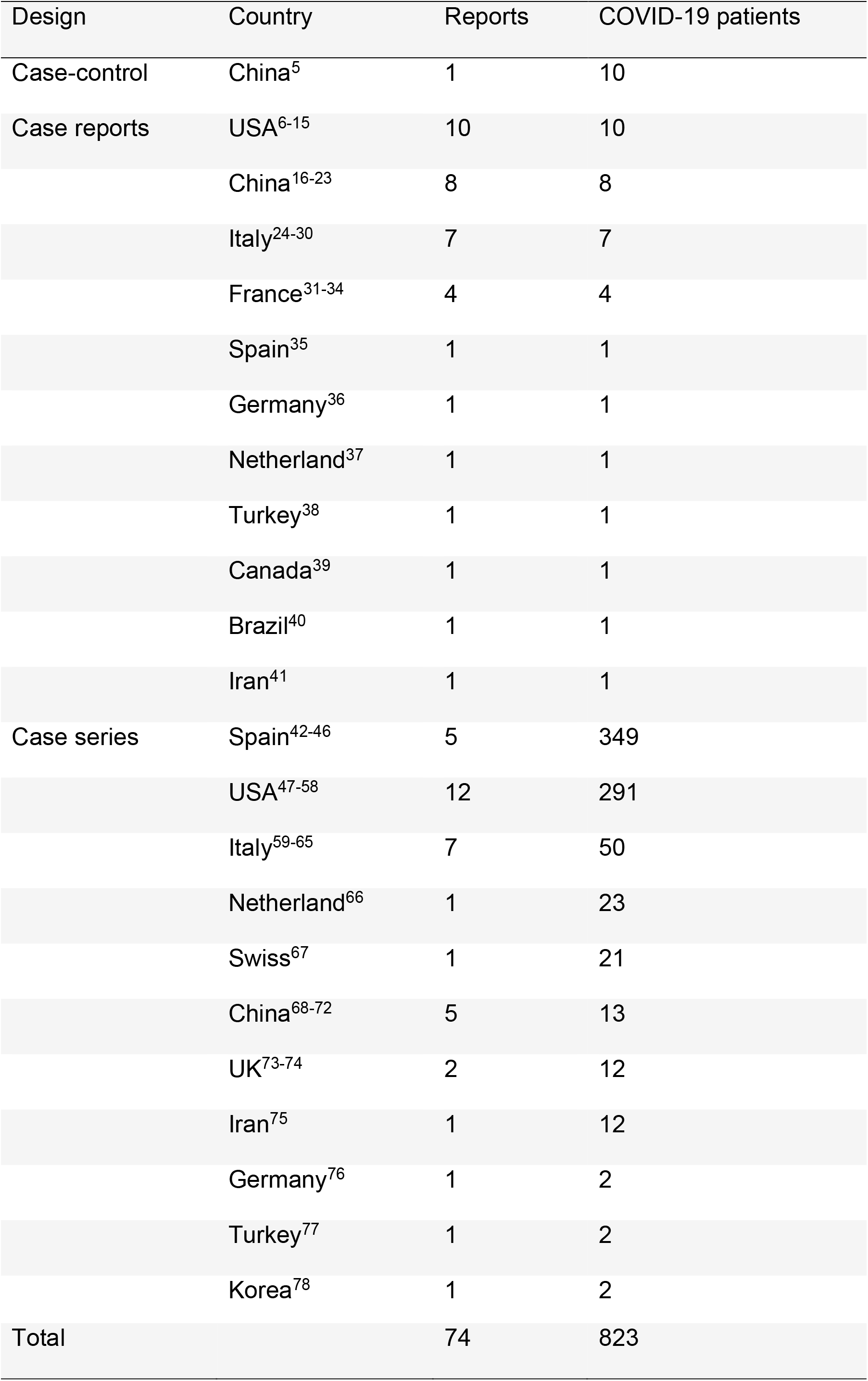
Distribution of the study designs of the review.

### Demographic data

Adult patients’ age ranged from 19-81 years. Seven pediatric patients – aged 6 months, 3 years, 4.5 years, 13 years, and 3 with unavailable age data – were also reported.

According to the available data, 554 (68.4%) male and 256 (31.6%) female patients, and 208 (75.6%) deceased-donor and 67 (24.4%) living-donor transplantation were reported.

Twenty-six cases developed COVID-19 within 1 year after transplantation (including 13 cases in the 1^st^ 3 months) and 111 cases after 1 year. Time since transplantation was not mentioned or detailed in 686 cases.

### Immunosuppression

Baseline immunosuppression could be only identified in 524 cases. Calcineurin inhibitors (CNI) were given in 463 cases (88.4%), mycophenolate mofetil (MMF) in 358 (68.3%), steroids in 313 (59.7%), mammalian target of rapamycin inhibitors (mTORi) in 40 (7.6%), and others (azathioprine, belatacept, or basiliximab) in about 6.3% of patients.

In response to COVID-19, immunosuppressive management differs between reports. CNI was reduced in 168 cases, discontinued in 91, maintained in 60, while newly started in 4 patients. MMF was discontinued in 203 cases, reduced in 34 cases, maintained in 21 cases, and started in 1 after COVID-19.

Patients started new steroids in 143 cases, continued or increased the dose of steroids in 104, discontinued in 8, and reduced the dose in 5. Adding or increasing the dose of steroids may be as a replacement of other immunosuppressants or as a treatment option of Acute Respiratory Distress Syndrome (ARDS). mTORi was discontinued in 14 patients, continued in 2, and reduced in one patient. (Table 2) summarizes the immunosuppressive management in COVID-19 transplant cases.

**Table 2.** Common Immunosuppressive therapy at baseline and changes after COVID-19 diagnosis.

| Drug | Baseline | Discontinued | Reduced | Maintained | Started |
| --- | --- | --- | --- | --- | --- |
| CNI (No., %) | 463/524<br>(88.4%) | 168 | 91 | 60 | 4 |
| MMF (No., %) | 358/524<br>(68.3%) | 203 | 34 | 21 | 1 |
| Steroids (No., %) | 313/524<br>(59.7 %) | 8 | 5 | 104* | 143 |
| mTORi (No., %) | 40/524<br>(7.6%) | 14 | 1 | 2 | - |

**CNI**, Calcineurin inhibitors, **MMF** mycophenolate mofetil, **mTORi** mammalian target rapamycin inhibitors
\*steroids maintained, increased in dose, or changed from oral to intravenous route

Thirty-five cases reported no immunosuppressive changes, and 36 cases reported a reduction of immunosuppression without details. Three-hundred and fifteen cases did not report immunosuppressive management after COVID-19.

### Clinical presentation, diagnosis, and case severity

The most common symptoms were fever (n= 577, 70%), cough (n= 520, 63%), dyspnea (n= 277, 33.7%), diarrhea (n= 153, 18.6%), myalgia (n= 105, 12.7%), and fatigue (n= 104, 12.6%). While, anorexia, loss of smell or taste, sore throat, nausea, and nasal congestion were infrequent (6.8%, collectively).

SARS-CoV-2 was confirmed with rt-PCR in 300 cases (36.5%). Chest x-ray and computed tomography (CT) scans showed abnormalities at the time of presentation in 255 (31%) and 97 (11.8%), respectively. Radiological findings were variable (bilateral or unilateral) in the form of ground-glass opacity, interstitial thickening, or infiltration. Lung ultrasound was available only in one case that showed progressive thick and confluent B lines, which improved with patient improvement.^9^ Sixty patients and twelve cases showed no abnormalities in chest x-ray and CT scans, respectively, at the time of presentation.

Laboratory results showed lymphopenia in 398/442 cases (90%), elevated C-reactive protein 213/302 (70.5%), elevated D-dimer 104/191 (54.5%), elevated ferritin 93/173 (53.8%), elevated troponin 71/132 (53.8%), elevated lactate dehydrogenase (LDH) 48/95 (50.5%), and elevated liver enzymes 25/158 (15.8%).

Among 372 patients, 114 (30.6%) were mild COVID-19, 101 (27.1%) moderate, and 157 (42.2%) severe or critical.

(Table 3) shows the distribution of case severity according to the transplanted organ.

**Table 3.** Distribution of illness severity and organ transplant.

| Organ | Mild | Moderate | Sever or Critical | Total |
| --- | --- | --- | --- | --- |
| Kidney | 44 | 35 | 90 | 169 |
| Liver | 26 | 7 | 29 | 62 |
| Heart | 11 | 1 | 11 | 23 |
| Lung | 4 | 1 | 9 | 14 |
| Combind | - | 2 | 2 | 4 |
| Non-specified | 29 | 55 | 16 | 100 |
| Total | 114 | 101 | 157 | 372 |

### Treatment

Four-hundred and sixty-nine patients (57%) received chloroquine or hydroxychloroquine, 170 (20.7%) ritonavir/lopinavir, 15 (1.8%) oseltamivir. Fifty-two (6.3%) patients received no antiviral therapy.

Two-hundred and eleven (25.6%) patients received prophylactic antibiotics, including 88 (10.7%) who received azithromycin – alone or in-combination with hydroxychloroquine.

Seventy-one patients received tocilizumab and 6 received leronlimab.

Oxygen therapy was reported in 212 patients (53 nasal cannulas, 20 non-invasive ventilation (NIV), 13 high-flow nasal cannula, and 126 non-specified) and 46 cases reported no oxygen therapy.

### Outcomes

One-hundred and twenty-one (14.7%) cases required ICU admission and 97 (11.8%) were mechanically ventilated.

Acute kidney injury (AKI) developed in 63 (7.7%) of cases. Twenty-two (35%) were kidney, 14 (22.2%) liver, 14 (22.2%) heart, 1 lung, and 12 non-specified organ transplant.

De novo dialysis started in 29 (3.5%) cases (14 kidney, 5 heart, 1 lung, and 9 non-specified organ transplant patients) and extracorporeal membrane oxygenation (ECMO) in 3 patients.

Mortality was reported in 160 (19.4%) of cases. None of the 7 pediatric patients in our cohort died. Of those 160 patients, 104 were kidney-transplant patients, 20 liver, 9 heart, 1 lung, and 26 non-specified organ-transplant patients.

## Discussion

The present scoping review identified 74 studies reporting 823 cases of COVID-19 infection in solid organ transplantation recipients. The common immunosuppressive practice was the dose reduction of CNI (52%), discontinuation of CNI (27.7%), discontinuation of MMF (78.7 %), discontinuation of mTORi (82.4%), and continuation (40%) or commencing (55%) steroids. Of 372 cases, COVID-19 was mild in 114 (30.6%) patients, moderate in 101 (27.1%), and severe or critical in 157 (42.2%). We found 121 (14.7%) ICU admissions, 97 invasive mechanical ventilation, and 63 (7.7%) with AKI. Mortality was reported in 160/823 (19.4%) of cases.

The variations in immunosuppressive therapy after COVID-19 reflect the theoretical controversies. Due to their immunosuppressed state, organ transplant recipients may be at higher risk for both being infected with SARS-CoV-2 and severe forms of COVID-19. On the other hand, immune suppression may ameliorate the cytokine surge, which is responsible for the systemic hyperinflammatory manifestations of COVID-19, including ARDS, multiorgan dysfunction syndrome, shock^79^, and the secondary hemophagocytic lymphohistiocytosis syndrome.^80^ Furthermore, immunosuppression reduction may cause graft rejection and immune constitution reaction causing paradoxical disease worsening.^80^ There are no solid recommendations regarding immunosuppressive protocol in transplant patients with COVID-19. There is even a recommendation against the routine reduction of CNIs.^79^

Generally, immunosuppressed patients present with atypical or attenuated signs and symptoms of infection, often leading to late presentations, worse outcomes, and prolonged virus shedding and infectivity.^24^

**Table 4** presents the common presentations of those patients versus non-transplant COVID-19 patients.^81^ Diarrhea was more frequent in transplant patients with COVID-19 (18.6% versus 3.8%); however, some reports showed a higher incidence of diarrhea in COVID-19 non-transplant cases.^82^ In a report of 90 organ transplant recipients, the incidence of diarrhea was even higher (31%).^48^ This may be attributed to MMF, which causes diarrhea.^83^ Diarrhea may be also associated with COVID-19 severity and the need for MV.^81^ Fatigue was less common in the transplant population (12.6% vs 38%). Dyspnea is the third common in both groups with a greater presence in transplant, 33.7 % vs 18.7% in non-transplant patients. In Covid-19 patients, dyspnea upon presentation is associated with a severe clinical course.^48^ This may be a marker of pulmonary disease severity in transplant recipients.

**Table 4.** Comparison between common clinical presentation and major outcomes of COVID-19 in transplant vs non-transplant patients.

|  | Transplant | Non-transplant<sup>81</sup> |
| --- | --- | --- |
| Fever | 70% | 88.7% |
| Cough | 63% | 67.8% |
| Dyspnea | 33.7% | 18.7% |
| Diarrhea | 18.6% | 3.8% |
| Myalgia/Arthralgia | 12.7% | 14.9% |
| Fatigue | 12.6% | 38% |
| ICU admission | 14.7% | 5% |
| Invasive ventilation | 11.8% | 2.3% |
| Acute kidney Injury | 7.7% | 0.5% |
| Mortality | 19.4% | 1.4% |

ICU, Intensive Care Unit
Data of the non-transplant patients are according to Guan et al.<sup>81</sup>

Among 442 available laboratory results, 90% of transplant patients showed lymphopenia vs 83.2% in the non-transplant population.^81^ Lymphopenia is associated with more ARDS, ICU admission, higher troponin, and more myocardial injury, and death.^84^

According to our findings, elevated CRP, D-dimer, ferritin, troponin, and LDH were higher in transplant patients when compared with the general population.^81^ Elevation of some or all of these parameters is associated with a severe COVID-19 course and a more unfavorable outcome. ^48,81^

Unfortunately, among our 823 cases, we could identify the disease severity of only 372 patients. Classification of severity was different between reports; some classified cases as mild, moderate, and critical, while others classified cases as mild, moderate, and severe.^5,48^ Therefore, we collected severe and critical cases in the same category as severe or critical illness. In our review, 42.2 % of reported patients were severe or critical, which is significantly higher when compared with the general population (15.7%).^81^ In a large non-transplant Chinese cohort, severe cases represented 14% and critical cases represented 5%.^85^ This copes with that immunosuppressed transplant recipients are at a higher risk for severe COVID-19 course.^79^

In a case-control study, one transplant patient died and transplant patients showed more severe course than non-transplant patients. The length of hospital stay and the duration of virus shedding (positive PCR) were longer in transplant cases. Therefore, transplant patients with COVID-19 might take a longer time to recover and remain infective for a longer duration.^5^

Nine (64.3%) of our 14 lung transplant patients were classified as severe or critical. Despite being a small cohort, being primarily a respiratory disease, COVID-19 may be more severe among lung transplant recipients.

Compared to non-transplant patients^81^, ICU admission was needed in 121 (14.7% versus 5%), invasive mechanical ventilation in 97 transplant cases (11.8% versus 2.3%),, and mortality in 160 (19.4% versus 1.4%) of cases.

On July 14, 2020, the combined SECURE-Cirrhosis and COVID-HEP registries reported ICU admission in 47/160 (29.4%), invasive ventilation in 31/160 (19.4%), and mortality in 30/160 (18.8%) in liver transplant recipients.^86^ Along with our results, these findings support more illness severity and fatality among transplant recipients with COVID-19.

Acute kidney injury (AKI) developed in 63 (7.7%) of cases including 29 patients who needed de novo dialysis. While AKI developed only in 0.5% of COVID-19 as found by Guan et al.^81^ This may be due to the more severe course of COVID-19 in transplant recipients, nephrotoxicity of some immune suppressants, and chronic rejection process in kidney transplant patients.

Forty-six hospitalized patients did not receive oxygen therapy and some reported no hospitalization for mild cases with favorable outcomes.^48^ Therefore, transplant patients with SARS-CoV-2 can be managed safely in the outpatient settings. This decision should be taken case-by-case keeping in mind the comorbidities and ability of rapid transfer to a transplant center in case of deterioration.^87^

Chloroquine, hydroxychloroquine, and various antiviral therapies were commonly used despite the lack of evidence and the known interactions with CNI.^87^

We had some limitations in our scoping review. This report only included case-reports, case series, and one case-control study; however, this was the best available data. Missed individual data and the aggregate data from the case series hindered us from extracting proper associations between case severity, immunosuppression protocol, and major outcomes. Our reporting was only up to the date of June 3, 2020. We did not include a critical appraisal of the primary reports. Patient duplication should be minimal as we traced cases carefully and contacted authors for primary source overlap.

In conclusion, our results suggest that COVID-19 in transplant patients has a more severe course, worse major outcomes (ICU admission, Invasive MV, AKI), and higher mortality than non-transplant patients.

## Supporting information

Search strategy

## Data Availability

Data will be available on request.

## Supporting information statement

Additional supporting information may be found online in the Supporting Information Section at the end of the article.

## Abbreviations

AKI: Acute Kidney Injury
ARDS: Acute Respiratory Distress Syndrome
CNI: Calcineurine inhibitor
CRP: C-reactive protein
CT: Computed Tomography
ECMO: Extracorporeal Membrane Oxygenation
ICU: Intensive Care Unit
LDH: Lactate Dehydrogenase
mTORi: mammalian Target Organ of Rapamycin inhibitor
MV: Mechanical Ventilation
MMF: Mycophenolate Mofetil
PRISMA: Preferred Reporting Items for Systematic Review and Meta-Analysis
rt-PCR: reverse transcription – Polymerase Chain Reaction
SARS-CoV-2: severe acute respiratory syndrome – coronavirus-2

## References

1. Fishman J. Infection in organ transplantation. American Journal of Transplantation. 2017;17(4):856–79.

2. Willicombe M, Thomas D, McAdoo S. COVID-19 and Calcineurin Inhibitors: Should They Get Left Out in the Storm? Journal of the American Society of Nephrology. 2020;31(6):1145–6.

3. Arksey H, O’Malley L. Scoping studies: Towards a methodological framework. Int J Soc Res Methodol. 2005;8:19–32.

4. Tricco AC, Lillie E, Zarin W, et al. PRISMA extension for scoping reviews (PRISMA-ScR): Checklist and explanation. Ann Intern Med. 2018;169:467–473.

5. Zhu L, Gong N, Liu B, et al. Coronavirus disease 2019 pneumonia in immunosuppressed renal transplant recipients: a summary of 10 confirmed cases in Wuhan, China. European Urology. 2020.

6. Hsu JJ, Gaynor P, Kamath M, et al. COVID-19 in a High-Risk Dual Heart and Kidney Transplant Recipient. American Journal of Transplantation. 2020.

7. Lagana SM, De Michele S, Lee MJ, et al. COVID-19 Associated Hepatitis Complicating Recent Living Donor Liver Transplantation. Archives of pathology & laboratory medicine. 2020.

8. Billah M, Santeusanio A, Delaney V, Cravedi P, Farouk SS. A catabolic state in a kidney transplant recipient with COVID-19. Transplant International. 2020.

9. Hammami MB, Garibaldi B, Shah P, et al. Clinical Course of COVID-19 in a Liver Transplant Recipient on Hemodialysis and Response to Tocilizumab Therapy: A Case Report. American Journal of Transplantation. 2020.

10. Johnson KM, Belfer JJ, Peterson GR, Boelkins MR, Dumkow LE. Managing COVID-19 in Renal Transplant Recipients: A Review of Recent Literature and Case Supporting Corticosteroid-sparing Immunosuppression. Pharmacotherapy: The Journal of Human Pharmacology and Drug Therapy. 2020;40(6):517–24.

11. Russell MR, Halnon NJ, Alejos JC, Salem MM, Reardon LC. COVID-19 in a pediatric heart transplant recipient: Emergence of donor-specific antibodies. The Journal of Heart and Lung Transplantation. 2020.

12. Bush R, Johns F, Acharya R, Upadhyay K. Mild COVID-19 in a pediatric renal transplant recipient. American Journal of Transplantation. 2020.

13. Allam SR, Dao A, Madhrira MM, et al. Interleukin-6 Receptor Antagonist Therapy to Treat SARS-CoV-2 Driven Inflammatory Syndrome in a Renal Transplant Recipient. Transplant Infectious Disease. 2020:e13326.

14. Kumar RN, Tanna SD, Shetty AA, Stosor V. COVID-19 in an HIV-positive Kidney Transplant Recipient. Transplant Infectious Disease. 2020:e13338.

15. Keller BC, L. A, Sobhanie M, et al. Early COVID-19 infection after lung transplantation. American Journal of Transplantation. 2020.

16. Qin J, Wang H, Qin X, et al. Perioperative presentation of COVID-19 disease in a liver transplant recipient. Hepatology. 2020.

17. Liu B, Wang Y, Zhao Y, Shi H, Zeng F, Chen Z. Successful treatment of severe COVID-19 pneumonia in a liver transplant recipient. American Journal of Transplantation. 2020.

18. Chen S, Yin Q, Shi H, et al. A familial cluster, including a kidney transplant recipient, of Coronavirus Disease 2019 (COVID-19) in Wuhan, China. American Journal of Transplantation. 2020.

19. Wang J, Li X, Cao G, Wu X, Wang Z, Yan T. COVID-19 in a kidney transplant patient. European Urology. 2020;77(6):769.

20. Ning L, Liu L, Li W, et al. Novel coronavirus (SARS-CoV-2) infection in a renal transplant recipient: case report. American Journal of Transplantation. 2020.

21. Huang JF, Zheng KI, George J, et al. Fatal outcome in a liver transplant recipient with COVID-19. American Journal of Transplantation. 2020.

22. Zhu L, Xu X, Ma K, et al. Successful recovery of COVID-19 pneumonia in a renal transplant recipient with long-term immunosuppression. American Journal of Transplantation. 2020.

23. Zhang M, Zhang J, Shi H, Liu B, Zeng F. Viral Shedding Prolongation in a Kidney Transplant Patient with COVID-19 Pneumonia. American Journal of Transplantation. 2020.

24. Bartiromo M, Borchi B, Botta A, et al. Threatening drug-drug interaction in a kidney transplant patient with Coronavirus Disease 2019 (COVID-19). Transplant Infectious Disease. 2020.

25. Seminari E, Colaneri M, Sambo M, et al. SARS Cov-2 infection in a renal-transplanted patient: A case report. American Journal of Transplantation. 2020.

26. Fontana F, Alfano G, Mori G, et al. Covid-19 pneumonia in a kidney transplant recipient successfully treated with Tocilizumab and Hydroxychloroquine. American Journal of Transplantation. 2020.

27. Gisondi P, Zaza G, Del Giglio M, Rossi M, Iacono V, Girolomoni G. Risk of hospitalization and death from COVID-19 infection in patients with chronic plaque psoriasis receiving a biologic treatment and renal transplant recipients in maintenance immunosuppressive treatment. Journal of the American Academy of Dermatology. 2020.

28. Lauterio A, De Carlis R, Belli L, Fumagalli R, De Carlis L. How to guarantee liver transplantation in the north of Italy during the COVID-19 pandemic: A sound transplant protection strategy. Transplant International. 2020.

29. Lauterio A, Valsecchi M, Santambrogio S, et al. Successful recovery from severe COVID-19 pneumonia after kidney transplantation: The interplay between immunosuppression and novel therapy including tocilizumab. Transplant Infectious Disease. 2020:e13334.

30. Bussalino E, De Maria A, Russo R, Paoletti E.Immunosuppressive therapy maintenance in a kidney transplant recipient with SARS-CoV-2 pneumonia: A case report. American Journal of Transplantation. 2020.

31. Marx D, Moulin B, Fafi-Kremer S, et al. First case of COVID-19 in a kidney transplant recipient treated with belatacept. American Journal of Transplantation. 2020.

32. Faguer S, Del Bello A, Abravanel F, Nicolau-Travers M-L, Kamar N. Tocilizumab for hemophagocytic syndrome in a kidney transplant recipient with COVID-19. Annals of Internal Medicine. 2020.

33. Morand A, Roquelaure B, Colson P, et al. Child with liver transplant recovers from COVID-19 infection. A case report. Archives de Pédiatrie. 2020.

34. Farfour E, Picard C, Beaumont L, et al. COVID-19 in lung-transplanted and cystic fibrosis patients: Be careful. Journal of Cystic Fibrosis. 2020.

35. Guillen E, Pineiro GJ, Revuelta I, et al. Case report of COVID-19 in a kidney transplant recipient: does immunosuppression alter the clinical presentation? American Journal of Transplantation. 2020.

36. Aigner C, Dittmer U, Kamler M, Collaud S, Taube C. COVID-19 in a lung transplant recipient. The Journal of Heart and Lung Transplantation. 2020;39(6):610.

37. Meziyerh S, Zwart TC, van Etten RW, et al. Severe COVID-19 in a renal transplant recipient: A focus on pharmacokinetics. American Journal of Transplantation. 2020.

38. Arpali E, Akyollu B, Yelken B, Tekin S, Turkmen A, Kocak B. Case report: A kidney transplant patient with mild COVID-19. Transplant Infectious Disease. 2020:e13296.

39. Xu JJ, Samaha D, Mondhe S, Massicotte-Azarniouch D, Knoll G, Ruzicka M. Renal Infarct in a COVID-19 Positive Kidney-Pancreas Transplant Recipient. American Journal of Transplantation. 2020.

40. de Barros Machado DJ, Ianhez LE. COVID-19 pneumonia in kidney transplant recipients-where we are? Transplant Infectious Disease. 2020:e13306.

41. Namazee N, Mahmoudi H, Afzal P, Ghaffari S. Novel Corona Virus 2019 pneumonia in a kidney transplant recipient. American Journal of Transplantation. 2020.

42. Fernández-Ruiz M, Andrés A, Loinaz C, et al. COVID-19 in solid organ transplant recipients: a single-center case series from Spain. American Journal of Transplantation. 2020.

43. Trujillo H, Caravaca-Fontán F, Sevillano Á, et al. SARS-CoV-2 Infection in Hospitalized Patients with Kidney Disease. Kidney International Reports. 2020;5(6):905–9.

44. Melgosa M, Madrid A, Alvárez O, et al. SARS-CoV-2 infection in Spanish children with chronic kidney pathologies. Pediatric Nephrology (Berlin, Germany). 2020:1.

45. Sánchez-Álvarez JE, Fontán MPs, Martín CJ, et al. Status of SARS-CoV-2 infection in patients on renal replacement therapy Report of the COVID-19 Registry of the Spanish Society of Nephrology (SEN). Nefrología (English Edition). 2020.

46. Crespo M, José Pérez-Sáez M, Redondo-Pachón D, et al. COVID-19 in elderly kidney transplant recipients. American Journal of Transplantation. 2020.

47. Program CUKT. Early description of coronavirus 2019 disease in kidney transplant recipients in New York. Journal of the American Society of Nephrology. 2020;31(6):1150–6.

48. Pereira MR, Mohan S, Cohen DJ, et al. COVID-19 in solid organ transplant recipients: Initial report from the US epicenter. American Journal of Transplantation. 2020.

49. Kates OS, Fisher CE. Earliest cases of coronavirus disease 2019 (COVID-19) identified in solid organ transplant recipients in the United States. 2020.

50. Akalin E, Azzi Y, Bartash R, et al. Covid-19 and Kidney Transplantation. The New England journal of medicine. 2020;382(25):2475–7.

51. Nair V, Jandovitz N. COVID-19 in kidney transplant recipients. 2020.

52. Holzhauser L, Lourenco L, Sarswat N, Kim G, Chung B, Nguyen AB. Early experience of COVID-19 in 2 heart transplant recipients: Case reports and review of treatment options. American journal of transplantation : official journal of the American Society of Transplantation and the American Society of Transplant Surgeons. 2020.

53. Husain SA, Dube G, Morris H, Fernandez H. Early Outcomes of Outpatient Management of Kidney Transplant Recipients with Coronavirus Disease 2019. 2020.

54. Ketcham SW, Adie SK, Malliett A, et al. Coronavirus Disease-2019 in Heart Transplant Recipients in Southeastern Michigan: A Case Series. Journal of cardiac failure. 2020;26(6):457–61.

55. Latif F, Farr MA, Clerkin KJ, et al. Characteristics and Outcomes of Recipients of Heart Transplant With Coronavirus Disease 2019. JAMA cardiology. 2020.

56. Lee BT, Perumalswami PV, Im GY, Florman S, Schiano TD. COVID-19 in Liver Transplant Recipients: An Initial Experience from the U.S. Epicenter. Gastroenterology. 2020.

57. Fung M, Chiu CY, DeVoe C, et al. Clinical Outcomes and Serologic Response in Solid Organ Transplant Recipients with COVID-19: A Case Series from the United States. American journal of transplantation : official journal of the American Society of Transplantation and the American Society of Transplant Surgeons. 2020.

58. Morillas JA, Marco Canosa F, Srinivas P, et al. Tocilizumab therapy in five solid and composite tissue transplant recipients with early ARDS due to SARS-CoV-2. American journal of transplantation : official journal of the American Society of Transplantation and the American Society of Transplant Surgeons. 2020.

59. Gandolfini I, Delsante M. COVID-19 in kidney transplant recipients. 2020.

60. Alberici F, Delbarba E, Manenti C, et al. Management Of Patients On Dialysis And With Kidney Transplant During SARS-COV-2 (COVID-19) Pandemic In Brescia, Italy. Kidney Int Rep. 2020;5(5):580–5.

61. Bhoori S, Rossi RE, Citterio D, Mazzaferro V. COVID-19 in long-term liver transplant patients: preliminary experience from an Italian transplant centre in Lombardy. The lancet Gastroenterology & hepatology. 2020;5(6):532–3.

62. Maggi U, De Carlis L. The impact of the COVID-19 outbreak on liver transplantation programs in Northern Italy. 2020.

63. Donato MF, Invernizzi F, Lampertico P, Rossi G. Health Status of Patients Who Underwent Liver Transplantation During the Coronavirus Outbreak at a Large Center in Milan, Italy. Clinical gastroenterology and hepatology : the official clinical practice journal of the American Gastroenterological Association. 2020.

64. Travi G, Rossotti R, Merli M, Sacco A, Perricone G, Lauterio A. Clinical outcome in solid organ transplant recipients with COVID-19: A single-center experience. 2020.

65. Cozzi E, Faccioli E, Marinello S, et al. COVID-19 pneumonia in lung transplant recipients: report of two cases. American Journal of Transplantation. 2020.

66. Hoek RA, Manintveld OC, Betjes MG, et al. Covid-19 in solid organ transplant recipients: A single center experience. Transplant International. 2020.

67. Tschopp J, L’Huillier A, Mombelli M, et al. First experience of SARS-CoV-2 infections in solid organ transplant recipients in the Swiss Transplant Cohort Study. American Journal of Transplantation. 2020.

68. Huang J, Lin H, Wu Y, et al. COVID-19 in posttransplant patients—report of 2 cases. American Journal of Transplantation. 2020.

69. Zhang H, Chen Y, Yuan Q, et al. Identification of kidney transplant recipients with coronavirus disease 2019. European Urology. 2020.

70. Zhong Z, Zhang Q, Xia H, et al. Clinical characteristics and immunosuppressant management of coronavirus disease 2019 in solid organ transplant recipients. American Journal of Transplantation. 2020.

71. Li F, Cai J, Dong N. First cases of COVID-19 in heart transplantation from China. The Journal of Heart and Lung Transplantation. 2020;39(5):496–7.

72. Cheng D, Wen J, Liu Z, Lv T, Chen J. Coronavirus disease 2019 in renal transplant recipients: report of two cases. Transplant Infectious Disease. 2020:e13329.

73. Banerjee D, Popoola J, Shah S, Ster IC, Quan V, Phanish M. COVID-19 infection in kidney transplant recipients. Kidney International. 2020.

74. Verma A, Khorsandi SE, Dolcet A, et al. Low prevalence and disease severity of COVID-19 in post liver transplant recipients–a single centre experience. Liver International. 2020.

75. Abrishami A, Samavat S, Behnam B, Arab-Ahmadi M, Nafar M, Taheri MS. Clinical Course, Imaging Features, and Outcomes of COVID-19 in Kidney Transplant Recipients. European Urology. 2020.

76. Koczulla RA, Sczepanski B, Koteczki A, et al. SARS-CoV-2 infection in two patients following recent lung transplantation. American Journal of Transplantation. 2020.

77. Kocak B, Arpali E, Akyollu B, et al., editors. A Case Report of Oligosymptomatic Kidney Transplant Patients with COVID-19: Do They Pose a Risk to Other Recipients? Transplantation Proceedings; 2020: Elsevier.

78. Kim Y, Kwon O, Paek JH, et al. Two distinct cases with COVID-19 in kidney transplant recipients. 2020.

79. Boettler T, Newsome PN, Mondelli MU, et al. Care of patients with liver disease during the COVID-19 pandemic: EASL-ESCMID position paper. JHEP Reports. 2020:100113.

80. Mehta P, McAuley DF, Brown M, et al. COVID-19: consider cytokine storm syndromes and immunosuppression. Lancet (London, England). 2020;395(10229):1033.

81. Guan W-j, Ni Z-y, Hu Y, et al. Clinical characteristics of coronavirus disease 2019 in China. New England journal of medicine. 2020;382(18):1708–20.

82. D’Amico F, Baumgart DC, Danese S, Peyrin-Biroulet L. Diarrhea during COVID-19 infection: pathogenesis, epidemiology, prevention and management. Clinical Gastroenterology and Hepatology. 2020.

83. Zhang Y, Ruiz P. Solid organ transplant-associated acute graft-versus-host disease. Archives of pathology & laboratory medicine. 2010;134(8):1220–4.

84. Terpos E, Ntanasis-Stathopoulos I. Hematological findings and complications of COVID-19. 2020;95(7):834–47.

85. Wu Z, McGoogan JM. Characteristics of and Important Lessons From the Coronavirus Disease 2019 (COVID-19) Outbreak in China: Summary of a Report of 72?314 Cases From the Chinese Center for Disease Control and Prevention. Jama. 2020.

86. SECURE-Cirrhosis, EASL COVID-HEP registries. combined weekly update 2020, July 14. Available from: https://covidcirrhosis.web.unc.edu/files/2020/07/Weekly-Data-Update-16-July-2020.png.

87. Gleeson SE, Formica RN, Marin EP. Outpatient Management of the kidney transplant recipient during the SARS-Cov-2 virus pandemic. Clinical Journal of the American Society of Nephrology. 2020;15(6):892–5.

