## Supplementary material for "COVID-19 in 823 Transplant patients: A Systematic Scoping Review": Search strategy

(S1) Search strategy:

PUBMED

(organ transplants[Mesh] OR transplant*[tw]) AND (coronavirus[Mesh] OR "covid 19"[nm] OR COVID-19[tw] OR COVID19[tw] OR coronavirus[tw]) NOT (animals[Mesh] NOT humans[Mesh])

CENTRAL

([mh organ transplants] OR transplant*:ti,ab,kw) AND ([mh coronavirus] OR "covid 19":kw OR COVID-19:ti,ab,kw OR COVID19:ti,ab,kw OR coronavirus:ti,ab,kw) NOT ([mh animals] NOT [mh humans])

Web of Science

(organ transplants OR transplant*) AND (coronavirus OR "covid 19" OR COVID-19 OR COVID19 OR coronavirus) NOT (animals NOT humans)

CINAHL

((MH "organ transplants+") OR transplant*) AND ((MH "coronavirus+") OR MW "covid 19" OR COVID-19 OR COVID19 OR coronavirus) NOT ((MH "animals+") NOT (MH "humans+"))

SCOPUS

((INDEXTERMS("organ transplants") OR TITLE-ABS-KEY("transplant*")) AND (INDEXTERMS("coronavirus") OR CHEM("covid 19") OR TITLE-ABS-KEY("COVID-19") OR TITLE-ABS-KEY("COVID19") OR TITLE-ABS-KEY("coronavirus")) NOT (INDEXTERMS("animals") NOT INDEXTERMS("humans"))
